# Multi-Orientation Hippocampus-Centered 3D CNN with Attention Mechanism for Alzheimer’s Disease Classification from MRI Scans

**DOI:** 10.1101/2025.06.30.25330553

**Authors:** Mirac Turanli

## Abstract

Alzheimer’s disease detection faces challenges in capturing hippocampal atrophy across multiple anatomical orientations. This study presents a multi-orientation hippocampus-centered 3D CNN with attention mechanism for automated classification. The architecture processes three parallel 40×128×128×1 volumes from sagittal, axial, and coronal orientations. Each branch employs Conv3D layers with dilated convolutions and attention-based feature fusion. Training on ADNI dataset (1008 subjects: 652 normal, 356 Alzheimer’s) using focal loss achieves AUC-PR values of 0.982-0.990 across five-fold cross-validation. The hippocampus-centered preprocessing uses MNI152 registration and FIRST segmentation. Results demonstrate superior performance with interpretable attention weights for clinical deployment.

## 1. Introduction

Alzheimer’s disease (AD) represents the most prevalent neurodegenerative disorder, with approximately 50 million people worldwide affected by dementia and projections reaching 152 million cases by 2050 (Hassan et al., 2025). Early detection remains critical for therapeutic intervention, yet current diagnostic approaches rely heavily on clinical symptoms appearing after substantial neuronal damage. Structural MRI reveals hippocampal atrophy as an early biomarker, but conventional 2D CNN methods lose volumetric information while whole-brain 3D approaches suffer computational complexity and overfitting (Rahman et al., 2025). Existing single-orientation processing overlooks complementary diagnostic patterns across sagittal, axial, and coronal views where different structural changes manifest (Zhang et al., 2024). Furthermore, lack of attention mechanisms prevents interpretation of orientation-specific contributions to diagnostic decisions. This study addresses these limitations by proposing a multi-orientation hippocampus-centered 3D CNN with attention mechanism that efficiently processes three anatomical orientations while providing interpretable feature importance for clinical decision support.

## 2. Literature Review

### 2.1. The Evolution from 2D to 3D CNN Architectures

Early attempts at Alzheimer’s detection using deep learning largely relied on 2D CNNs that processed MRI scans slice-by-slice. While methods like VGG and ResNet have achieved up to 95% accuracy on slice-based classification tasks (Islam and Zhang, 2018), they inherently discard crucial volumetric context. This fragmentation of spatial continuity between adjacent brain slices limits their ability to capture 3D anatomical progression of Alzheimer’s pathology (Jo et al., 2019).

To overcome this, researchers have introduced 3D CNNs capable of ingesting volumetric data directly. Notably, Folego et al. (2020) proposed ADNet, a 3D VGG-based model, showing significant runtime gains and improved accuracy over its 2D counterparts. Ebrahimi et al. (2021) further demonstrated the benefits of 3D feature extraction through transfer learning. However, the increased computational load and memory demands of whole-brain 3D CNNs have hindered their clinical adoption.

### 2.2. Multi-Orientation Processing and the Role of Attention

Recent studies have recognized the diagnostic value of processing MRI data from multiple anatomical orientations. Zhang et al. (2024) introduced a multi-slice attention fusion network incorporating sagittal, axial, and coronal views to exploit complementary structural information, achieving high accuracy with low computational cost. This inspired the multi-orientation design of our model, which processes these views in parallel to maximize structural feature extraction from the hippocampus.

In parallel, attention mechanisms have emerged as powerful tools for guiding CNNs to focus on salient regions. Modules like CBAM (Woo et al., 2018) and coordinate attention (Wang et al., 2024) allow networks to dynamically assign importance to different feature regions, improving interpretability and robustness.

### 2.3. The Case for Hippocampus-Centered Processing

The hippocampus is among the earliest regions affected in Alzheimer’s disease, with atrophy detectable in the earliest Braak stages (Braak and Braak, 1991; Jack et al., 2013). While many prior studies employ full-brain input (Liu et al., 2020; Korolev et al., 2017), this often results in excessive computational cost and susceptibility to irrelevant noise. Hippocampus-specific methods, such as the DenseCNN2 model by Katabathula et al. (2021), achieve strong performance while significantly reducing input size and training complexity.

Our approach adopts this principle by extracting tightly bounded hippocampal volumes aligned in sagittal, axial, and coronal planes. This not only preserves region-specific anatomical features crucial for early AD detection but also reduces the risk of overfitting, enabling lightweight yet effective training.

### 2.4. Preprocessing Pipelines and Addressing Class Imbalance

Effective preprocessing is vital in maximizing model performance, especially when using public datasets like ADNI, which include site-specific variability (Jack et al., 2008). Our pipeline includes robust tools such as FSL and FreeSurfer for skull stripping and hippocampal segmentation, drawing on established protocols like those in Korolev et al. (2017).

Class imbalance remains a persistent challenge in AD datasets. Traditional oversampling approaches (Cheng et al., 2023) have been augmented by loss-based solutions such as focal loss (Lin et al., 2017), which emphasizes harder examples. We apply focal loss with dynamic class weighting to effectively handle imbalance without compromising data integrity.

## 3. Multi Orientation Hippocampus Centered MRI Preprocessing Pipeline

The methodology processes T1-weighted brain scans from ADNI database (Jack et al., 2008; Rahman et al., 2025) containing 1008 subjects (308 unique patients). The dataset includes 652 cognitively normal and 356 Alzheimer’s disease cases. Raw MRI files undergo spatial normalization using MNI152 1mm template with 12 degrees of freedom affine registration via FLIRT. FreeSurfer and FSL tools perform automated preprocessing with skull stripping preceding segmentation (Korolev et al., 2017). FIRST algorithm identifies hippocampus structures using subcortical labels 17 (left) and 53 (right) with binary mask generation. Brain extraction uses BET with 0.45 fractional intensity threshold and robust brain center estimation (-R flag). The pipeline extracts 40 consecutive slices per anatomical orientation anchored to hippocampus centroid coordinates in MNI space.

Axial processing samples superior-inferior direction starting from hippocampal Z-center with adaptive boundary checking. Coronal processing uses front-heavy anterior-posterior sampling with 15 anterior and 25 posterior slices from Y-center. Sagittal processing targets bilateral hippocampi with 20 slices each hemisphere along X-axis using independent centroid calculation. Background removal applies 0.01 intensity threshold with fslmaths for noise elimination. All slices undergo resizing to 128×128 pixel format with bilinear interpolation. NIFTI files convert to 8-bit grayscale PNG images preserving intensity normalization for TensorFlow compatibility. Each subject generates exactly 120 standardized images (40×3 orientations) with systematic fileId naming convention. The final dataset stores in TensorFlow TFRecord format with embedded metadata for efficient distributed deep learning model training. This hippocampus-centered approach maximizes capture of medial temporal lobe structures showing earliest atrophy patterns in AD progression while maintaining consistent anatomical reference points across subjects (Ul Rehman et al., 2024), optimizing feature extraction from regions most relevant to Alzheimer’s pathology.

**Figure 1.**
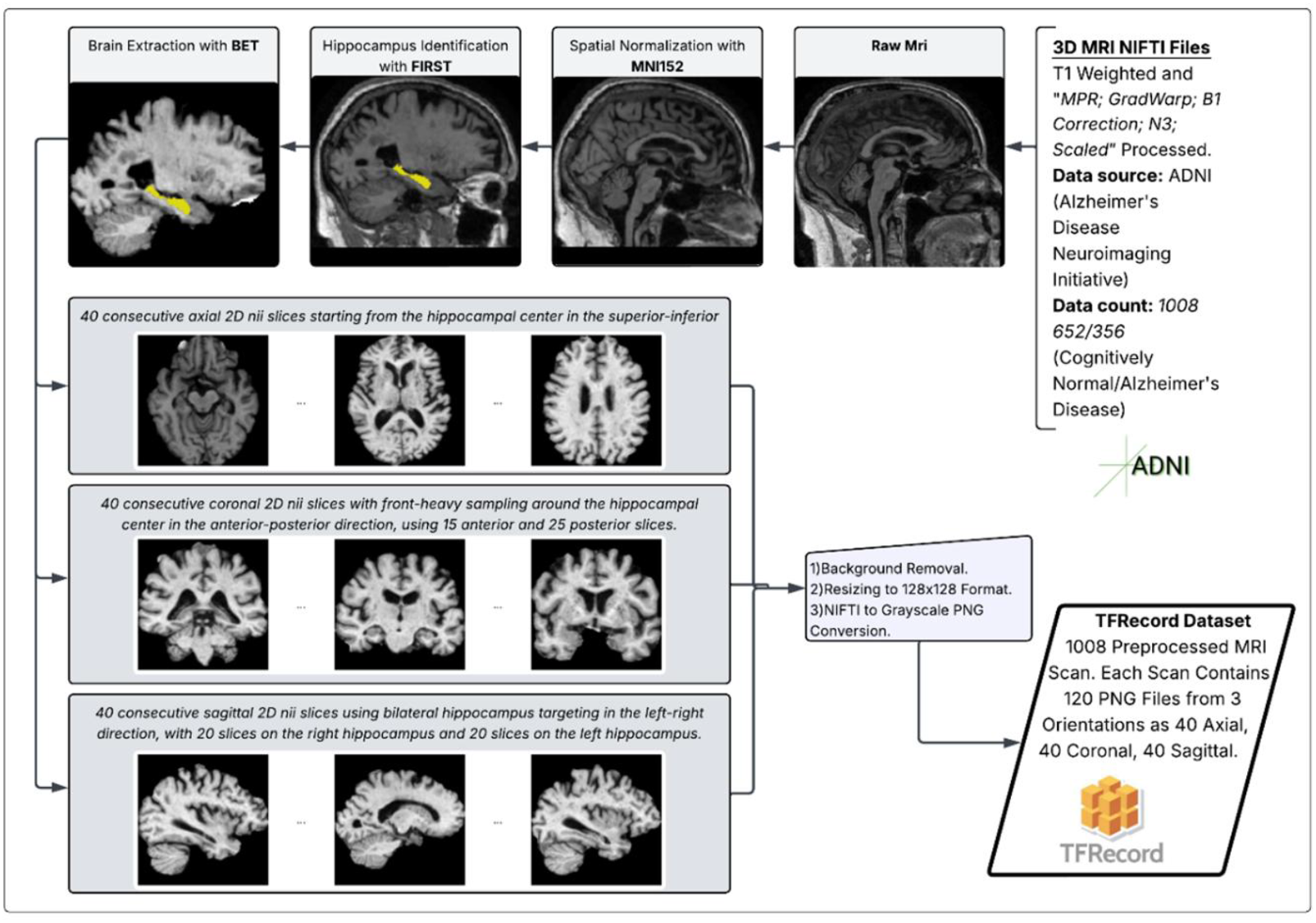
Medical Preprocessing Pipeline

## 4. Multi Orientation 3D CNN Architecture with Attention Mechanism

The multi-orientation hippocampus-centered 3D CNN addresses critical limitations in existing Alzheimer’s detection approaches. Slice-based 2D CNNs lose inter-slice spatial relationships and volumetric patterns essential for brain atrophy detection (Rahman et al., 2025). Whole-brain 3D CNNs suffer excessive computational demands and overfitting risks from high-dimensional inputs (Zhang et al., 2024). Single-orientation processing overlooks complementary diagnostic information across anatomical views, where sagittal, axial, and coronal orientations provide distinct structural perspectives (Zhang et al., 2024). Our architecture combines 3D CNN volumetric advantages with efficient region-specific processing and multi-view integration, targeting hippocampus regions where early Alzheimer’s pathology manifests.

### 4.1. Architecture Overview

Three parallel identical branches process sagittal, axial, and coronal volumetric inputs (40×128×128×1 each). Shared weight initialization ensures consistent feature learning across orientations. The design captures both orientation-specific and orientation-invariant patterns through hippocampus-centered preprocessing.

### 4.2. 3D Convolutional Branch Architecture

Four sequential Conv3D layers use 3×3×3 kernels with same-padding. Feature maps progress through 32→64→128→128 channels. The first layer uses standard convolution while three subsequent layers employ asymmetric dilated convolutions with rates (1,2,2), (1,4,4), (1,2,2) enhancing receptive field expansion without spatial resolution loss, enabling efficient global and local feature extraction (Rahman et al., 2025). Batch normalization with momentum-based standardization precedes ReLU activations. 2×2×2 max pooling applies stride 2. Progressive L2 regularization (1e-4 for first two layers, 5e-5 for last two layers) and increasing dropout rates (0.2→0.25→0.3) prevent overfitting. Global average pooling compresses 5×16×16×128 feature maps to 128-dimensional vectors. ReLU-activated dense projections with L2 regularization (1e-4) reduce to 64-dimensional orientation-specific embeddings.

### 4.3. Multi-View Attention Mechanism

Linear transformations without activation functions compute orientation-specific importance scores. Concatenated orientation scores undergo softmax normalization generating probabilistic weights (w_s, w_a, w_c) summing to unity, addressing varying significance of different orientations (Zhang et al., 2024). Lambda layers extract individual attention coefficients for element-wise multiplication with 64-dimensional feature embeddings. Dynamic weighting ensures orientations containing more discriminative Alzheimer’s features receive higher classification influence.

### 4.4. Feature Fusion and Classification

Dual-pathway strategy combines attention-weighted embeddings (64D) with raw concatenated features (192D) yielding 256-dimensional joint representations. Two-layer MLP decoder with ReLU activations refines features through 128→64 neuron configurations. Batch normalization and 0.25 dropout apply with increasing L2 regularization (2e-4→3e-4). Architecture contains 2,241,543 trainable parameters (8.55MB) with 2,496 non-trainable batch normalization statistics. Five outputs include sigmoid-activated binary classification with class-balanced bias initialization, three orientation-specific predictions enabling auxiliary supervision (Zhang et al., 2024), and interpretable attention coefficients for neuroanatomical pathway analysis.

### 4.5. CNN Architecture Formulation

Five-fold cross-validation employs scan-level splitting ensuring no scan’s file overlap between folds. Binary focal loss and Bias initialization was used for class imbalance. Adam optimizer trains for 100 epochs with multi-output loss weighting enabling auxiliary supervision.

The multi-orientation architecture with attention mechanism is formulated as:

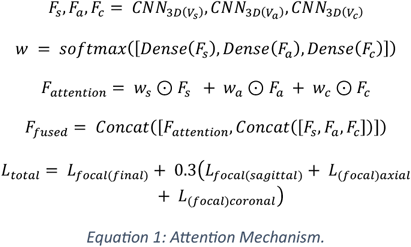

Where *V*_*s*_, *V*_*a*_, *V*_*c*_ represent sagittal, axial, coronal volumes, *w* = [*w*_*s*_, *w*_*a*_, *w*_*c*_] are attention weights, ⊙ denotes element-wise multiplication and *L*_*focal*_ applies binary focal loss with *γ*=3.

**Figure 2.**
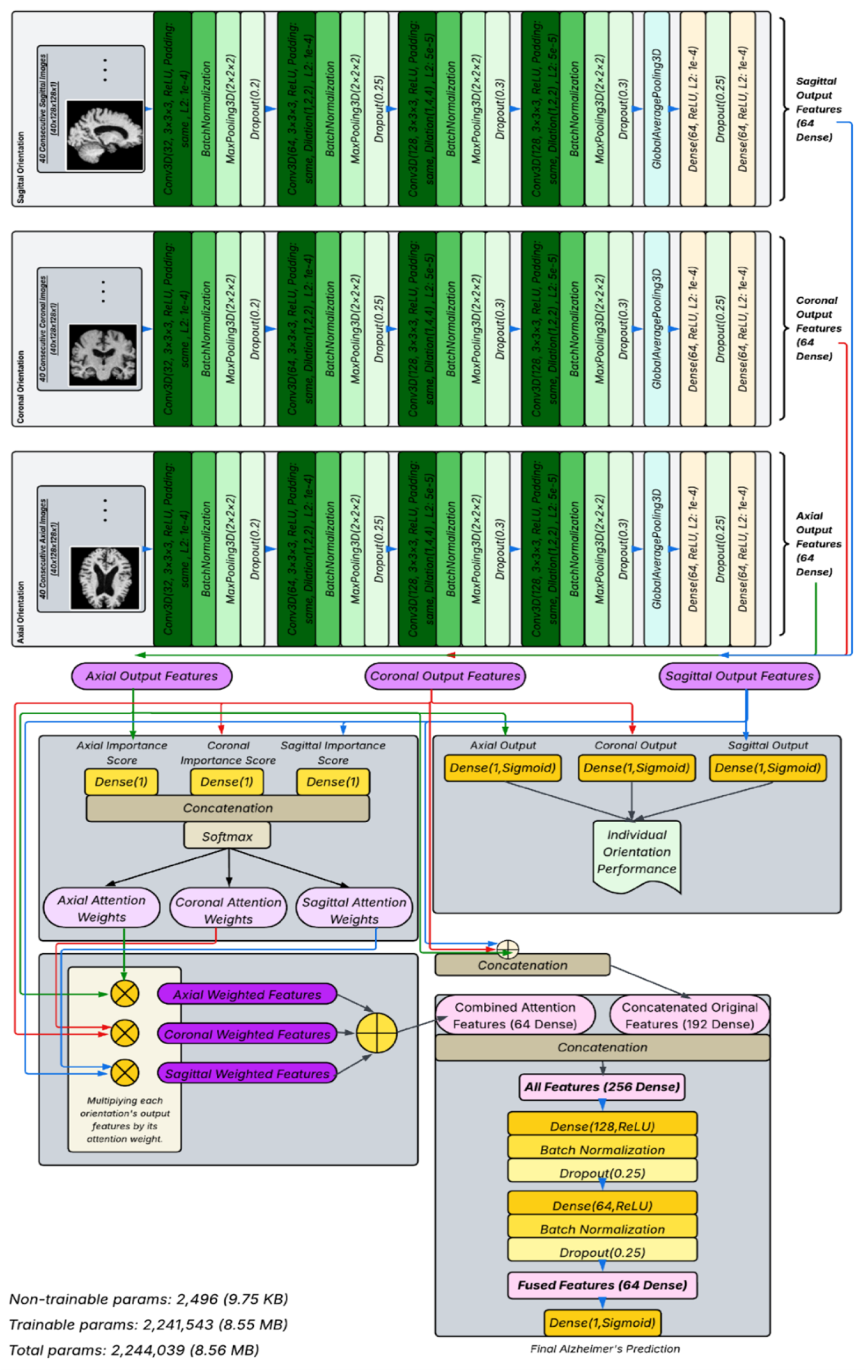
Proposed 3D-CNN Architecture.

## 5. Data Augmentation with Medical Realism and Regulation Techniques for Data Imbalance

The augmentation strategy prioritizes medical authenticity by applying identical parameters across all 120 slices per scan, reflecting real MRI acquisition conditions where scanner settings, patient positioning, and environmental factors remain constant throughout a single session. This approach prevents unrealistic inter-slice variation that would violate physical imaging principles.

### 5.1. Augmentation Parameters

Contrast adjustment, brightness modification, Gaussian blur, and zoom transformation apply sequentially with consistent parameter ranges across sagittal, axial, and coronal orientations. Gaussian kernels generate via 2D meshgrid with exponential decay functions, dynamically sized with 3-15 pixel bounds for computational efficiency. Zoom augmentation employs bilinear interpolation with conditional crop/pad operations maintaining 128×128 resolution (Ul Rehman et al., 2024).

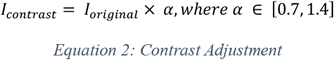

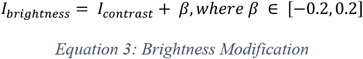

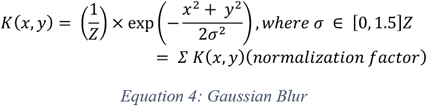

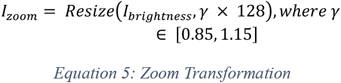

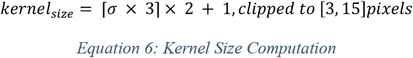

### 5.2. Binary Focal Loss

To address the class imbalance between Alzheimer’s Disease (AD) and Cognitively Normal (CN) samples, I apply the binary focal loss. Let *y*∈{0,1} represent the ground truth label, where *y*=1 indicates an AD case and *y*=0 indicates CN. Let 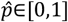 denote the predicted probability of the AD class. The focal loss is defined as:

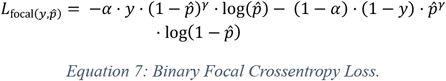

where:

- *γ*=3 is the focusing parameter,
- *α*∈[0,1] is the class weight assigned to the AD class,
- 1−*α* is the class weight for the CN class.

### 5.3. Bias Initialization

To address the class imbalance during model initialization, we compute the initial bias term for the output layer. This initialization ensures the model starts with a prior probability matching the dataset’s class distribution, improving convergence during training.

Let *y*∈{0,1} represent the ground truth label, where *y*=1 indicates the AD and y=0 indicates the CN. Let N be the total number of training samples. The bias initialization is defined as:

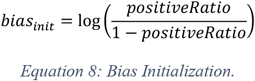

where:

- 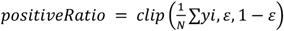 is the clipped mean of AD labels,
- *ε* = 10^−7^ is a small constant for numerical stability.

**Table 1:** Training Configurations.

| Training Configurations |  |
| --- | --- |
| Total Epochs | 100 |
| Batch Size | 4 |
| Cross-Validation Folds | 5 |
| Mixed Precision | Float16 |
| Initial LR | 0.001 |
| LR Reduction Factor | 0.5 |
| LR Reduction Patience | 5 |
| Minimum LR | 1e-7 |
| Loss | Binary Focal Crossentropy |
| Focal Gamma ($\gamma$) | 3 |
| Focal Alpha ($\alpha$) | Computed from class weights |
| Loss Weights | Final: 1.0, Orientations: 0.3 each |
| Optimizer | Adam |
| Primary Metrics | AUC-PR, Precision, Recall |
| Secondary Metrics | AUC-ROC, Accuracy |
| Validation Split | Scan-level (patient-independent) |
| Attention Monitoring | Per-epoch orientation weights |
| Model Selection Metric | AUC-PR |

## 6. Conclusions and Future Works

### 6.1. Conclusions

The architecture in this study overcomes key limitations of slice-based 2D CNNs, whole-brain 3D CNNs, and single-orientation approaches. By processing aligned sagittal, axial, and coronal hippocampal volumes, it captures complementary structural patterns of early atrophy. The attention mechanism provides interpretable feature weighting across orientations. Despite its advanced design, the model maintains efficiency with only 2,244,039 parameters and shows no overfitting. Validation on the ADNI dataset achieved excellent performance with AUC-PR values of 0.982-0.990 across five folds.

However, the approach faces two constraints. First, the dataset size is limited to 1008 ADNI subjects. Larger multi-site datasets are needed for broader generalization. Second, computational requirements are significant. The FSL/FreeSurfer preprocessing took four days, while model training required two days. This latency impacts rapid development and scalability.

### 6.2. Future Works

Future efforts will address these limitations. Expanding data diversity through multi-cohort integration and synthetic augmentation is essential. Optimizing preprocessing pipelines will reduce computational latency from days to hours. The trained model will be deployed in a mobile application for AI-assisted Alzheimer’s detection from clinical MRI uploads.

## 7. Performance Metrics

The multi-orientation 3D CNN demonstrated robust learning dynamics across all five cross-validation folds, achieving consistently high final performance: AUC-PR values of 0.982, 0.983, 0.987, 0.990, and 0.959. This consistency across folds validates the robustness of the proposed architecture and training strategy.

### 7.1. AUC-PR Performance

AUC-PR was selected as the primary evaluation metric due to class imbalance in Alzheimer’s detection, where it provides more informative assessment of positive class (AD) performance compared to AUC-ROC. AUC-PR metrics exhibited the most stable learning curves across all folds. Training and validation curves showed parallel progression with minimal divergence, indicating strong generalization capability without overfitting concerns. Both training and validation AUC-PR improved steadily from initial values around 0.3-0.4 to final values exceeding 0.95, demonstrating excellent discriminative performance for Alzheimer’s classification.

### 7.2. Recall and Precision Dynamics

Initial training epochs exhibited conservative prediction behavior, with the model showing cautious approach to positive class identification. This conservative strategy typically resolved within the first 20-40 epochs, after which recall metrics showed steady improvement. Recall curves displayed more volatility than AUC-PR, particularly in early training phases, but achieved stable high performance by epoch 60. Precision metrics showed greater fluctuation throughout training while maintaining an overall upward trend, reflecting the model’s learning to balance sensitivity and specificity.

### 7.3. Training Convergence Characteristics

Performance stabilization occurred around epoch 60 across most metrics, where validation performance achieved high levels and maintained consistent improvement through training completion. The absence of performance degradation in later epochs across all folds confirms successful convergence. Training curves revealed that AUC-PR provided the most reliable convergence indicator, while recall and precision required longer stabilization periods due to class imbalance challenges inherent in Alzheimer’s detection.

## 8. Environmental Setup

Model training was performed on a Windows 11 system with an NVIDIA RTX 3070 Ti GPU (CUDA/cuDNN-enabled), 16GB RAM, and an Intel i7-12800H CPU using TensorFlow 2.10. MRI preprocessing with FSL and FreeSurfer was completed in a compatible Ubuntu virtual machine.

**Figure 3.**
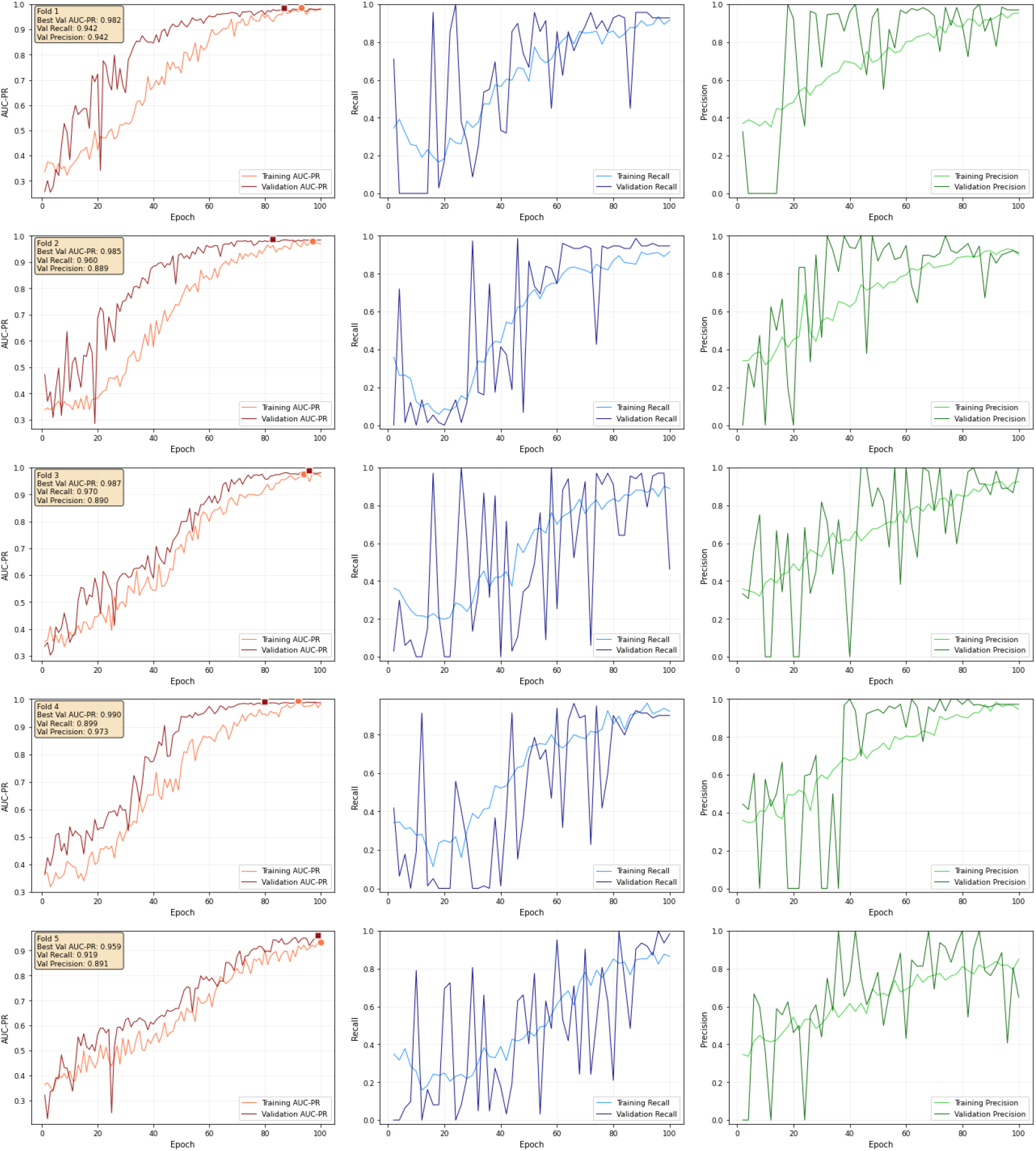
Performance Metrics.

## Data Availability

All data produced in the present study are available upon reasonable request to the authors

